# Social support for self-care: patient strategies for managing diabetes and hypertension in rural Uganda

**DOI:** 10.1101/2020.10.01.20205138

**Authors:** Andrew K. Tusubira, Christine K Nalwadda, Ann R Akiteng, Evelyn Hsieh, Christine Ngaruiya, Tracy L Rabin, Nicola Hawley, Robert Kalyesubula, Isaac Ssinabulya, Jeremy I Schwartz, Mari Armstrong-Hough

## Abstract

**Background:** The growing burden of non-communicable diseases (NCDs) threatens low-income countries. Self-care practices are crucial for successful management of NCDs to prevent complications. We sought to understand self-care efforts and their facilitators among patients with diabetes and hypertension in rural Uganda.

**Methods:** Between April and June 2019, we conducted a cross-sectional qualitative study among adult patients from outpatient NCD clinics at three health facilities in Uganda. We conducted 19 in-depth interviews exploring treatment practices and response to symptoms. We used content analysis to identify emergent themes.

**Results:** Three themes emerged in patients’ descriptions of their self-care practices. First, patients preferred conventional medicines as their first line of resort, but often used traditional medicines. In particular, patients used traditional medicines to mitigate the negative impacts of inconsistent access to conventional medicines and to supplement those medicines. Second, patients adopted a wide range of vernacular practices to supplement treatments and unavailable diagnostic tests, including tasting their urine to gauge blood-sugar level. Finally, patients sought social support for self-care activities, relying on networks of family members and peers for instrumental and emotional support. Patients saw their children as the most reliable source of support, especially money for medicines, transport and home necessities.

**Interpretation:** Patients valued conventional medicines but also engaged in varied self-care practices. They depended upon networks of social support from family and peers to maintain self-care. Interventions to improve self-care may be more effective if they improve access to medicines and engage or enhance patients’ social support networks.

## Background

Non-communicable diseases (NCDs) now account for the majority of the global burden of disease, and that burden is disproportionately higher in low- and middle-income countries (LMIC).^1, 2^ NCDs threaten resource-limited countries even as they continue to contend with a substantial burden of infectious diseases.^1, 3^ Studies from Malawi and Uganda show that the economic burden of chronic NCDs is particularly high for individuals and families in rural settings, where patients lack resources to access and maintain treatment.^4-6^ LMICs must therefore identify cost-effective approaches to manage their growing burden of NCDs. One such approach is to strengthen self-care practices among those living with NCDs.

Self-care is a patient-driven decision-making process regarding “behaviors that maintain physiologic stability and the response to symptoms when they occur”.^7^ Self-care includes all actions patients take to attain and maintain good health.^8^ Conventionally recommended self-care practices for patients with hypertension or diabetes include a diet rich in fruits and vegetables, adherence to medication, regular physical activity, smoking cessation, weight loss, and moderation of alcohol consumption.^9^ These practices can significantly improve health outcomes for patients with NCDs, reducing the costs of care.^10^

Social support is an important driver of self-care for diabetes and hypertension.^11^ Social support may take the form of physical, financial, or psychological help from family, friends, and community members.^12^ It may consist either of emotional support (i.e., a confidant) or instrumental support (i.e., tangible and/or physical assistance). For patients with chronic diseases, such support builds and sustains resilience, confidence to perform, and confidence to maintain appropriate self-care practices.^13^

However, little is known about the self-care practices of patients living with diabetes and/or hypertension in LMIC. Moreover, the relationship of care practices to patients’ social and cultural context, is under-addressed in the literature. We therefore explored how patients engaged in self-care practices for diabetes and hypertension in a rural Ugandan setting. In Uganda, the prevalence of diabetes and hypertension is increasing and most persons living with diabetes and/or hypertension experience suboptimal glycemic and/or blood pressure control.^14-16^ Persons living with NCDs in Uganda have been observed to seek varied channels of care and exhibit poor adherence to conventional treatment regimens.^5^

## Methods

### Study design

We carried out qualitative in-depth interviews with patients in rural Uganda who had been diagnosed with diabetes and/or hypertension to explore their practices to attain and maintain health.

### Study setting

This study was set at three health facilities in Nakaseke District, a rural district in central Uganda. Nakaseke has Uganda’s highest hypertension prevalence at 28.5% among adults.^14^ Nakaseke General Hospital, Semuto Health Center-IV, and Nakaseke LifeCare Medical Center host specialized NCD clinics for diabetes and hypertension on different days of the week. On average, Nakaseke General Hospital serves 75 patients, Semuto 10 patients, and LifeCare Center 15 patients at their NCD clinics each week. There, a nurse or clinician checks weight, blood pressure, and blood sugar; delivers health education; prescribes medications; and schedules monthly follow-up appointments. Medications are dispensed at a clinic-based pharmacy.

### Study population

We included patients above 18 years of age who 1) attended outpatient NCD clinics at one of the three selected facilities and 2) had been diagnosed with diabetes and/or hypertension at least three months prior to data collection.

### Sampling and sample size

We purposively selected interview participants for maximum variation by condition and gender. We selected at least two patients with hypertension, two with diabetes and two with both conditions at each health facility. We selected at least one male and one female for each condition category. A total of 19 patients, with at least six from each health facility, were invited to interview. We initially selected 18 participants. During initial interviews, we learned about the phenomenon of patient group leaders. We therefore selected the 19^th^ participant in order to include at least one patient leader. Research assistants approached patients in the waiting area to inform them about the study prior to receiving care. After the patient completed their visit, the selected participants were invited to complete the interview.

### Data collection

Three (one male and two female) trained and experienced research assistants with backgrounds in social science carried out in-depth interviews using a semi-structured interview guide. The guide was developed and reviewed by the research team and pretested among four patients with diabetes and hypertension attending care at Kasangati Health Center IV. After pretesting, we revised study instruments to ensure that questions were clear, appropriate, and aligned with the overarching research question.

The interview guide (Appendix 1) included five key domains: 1) perspectives on their condition(s), 2) experiences receiving medical care, 3) challenges to medication adherence, 4) symptom recognition and responses, and 5) other practices intended to prevent adverse conditions or to be well. All interviews were conducted in-person in the local language, Luganda, and lasted 30 to 45 minutes. Participants were compensated 15,000/= Ugandan Shillings for participation. Audio recordings were transcribed verbatim in Luganda and later translated to English. All data were collected between April and June, 2019.

### Analysis

We used a conventional approach to thematic content analysis to inductively derive categories.^17^. A team of four researchers, including AKT and MAH, independently read and coded three transcripts to develop initial codes. Coders discussed and reached consensus about the initial code structure, then applied it to two new transcripts while remaining open to new codes. Another meeting was held to further develop the code structure. The resulting structure was applied to all transcripts using Atlas.ti version 8.5. Emerging findings were refined through peer debriefing and works-in-progress sessions as part of a training grant (NIH D43 TW009607-07) and review with co-investigators. We present representative quotations for each theme.

### Human subjects and ethics approval

We received approval from the Makerere University School of Medicine Ethics Review Committee, the Yale Human Subjects Committee, and the Uganda National Council of Science and Technology (HS 2698). Administrative clearance was obtained from health facilities. All study participants provided written informed consent.

## Results

We enrolled all 19 patients invited to participate (Table 1). Their mean age was 55 years (standard deviation ±12 years). Most were farmers (n=10, 53%) and had attained at least primary-level formal education (n=17, 89%). Six (32%) participants were living with hypertension, six (32%) diabetes, and seven (36%) both conditions.

**Table 1:** Patient participant characteristics.

| **Characteristic** | **Frequency** | **Percent** |
| --- | --- | --- |
| **Age groups** |  |  |
| 35 – 49 | 5 | 26% |
| 50 – 59 | 8 | 42% |
| 60 and above | 6 | 32% |
| **Gender** |  |  |
| Male | 10 | 53% |
| Female | 9 | 47% |
| **Education** |  |  |
| No education | 2 | 11% |
| Primary | 8 | 42% |
| Secondary | 7 | 37% |
| Tertiary | 2 | 11% |
| **Occupation** |  |  |
| Peasant farmer | 10 | 53% |
| Business | 5 | 26% |
| Other | 4 | 21% |

Three overarching themes emerged. First, patients preferred conventional medicines as their first line of resort, but often used traditional medicines. Second, patients adopted a wide range of vernacular practices to supplement treatment. Finally, patients relied on both emotional and instrumental social support from family members and peers to mitigate uncertain access to medicines.

### Patients preferred conventional medicines but often used traditional medicines

Patients reported that clinician-prescribed conventional medicines were their key resource for self-care. However, their ability to adhere to the prescribed regimen was affected by inconsistent access to prescribed medicines. A 40-year-old woman explained:

> *In order to survive, I have to do what they [clinicians] told me and remain on medication. But the main challenge we have is getting medicines*.*… So I sometimes miss taking the medicines*. (IDI-8, HTN & DM, Female, 40)

Because they lacked consistent access to inexpensive medicines from health facility pharmacies, patients said they had to “improvise”:

> *I adhere to them and I take my medication as prescribed by the doctor, only that the medicines we receive are not enough for a month. We just improvise and buy in order to maintain our health. They* [clinicians] *normally tell us to go and buy the medicines, yet the medicines are very expensive*. (IDI-4, HTN&DM, Male, 65)

Because of insufficient availability in public pharmacies, patients reported being dispensed fewer doses than prescribed. Although patients sought to buy medicines at private pharmacies, they typically secured fewer than the prescribed number of doses.

When unable to access medicines at public facilities or afford medicines at private pharmacies, patients sought other remedies. Participants reported using traditional remedies, including local herbs, bitter vegetables, and salt water, in place of unavailable prescribed medicines (Table 2A). They saw herbal remedies as a stopgap when unable to afford prescribed medicines:

> ***I: Are there things you do when you have failed to get the recommended dose?***
>
> *R: Yes, I use herbals such as stem pieces of bitter trees. I boil them and drink. I also get kasaana* (Acacia hockii) *tree roots, which I boil and drink… there is a time I failed to raise money and resorted to using these herbs in order to prevent getting an attack due to this disease*. (IDI-10, HTN & DM, Female, 59)

**Table 2A:**
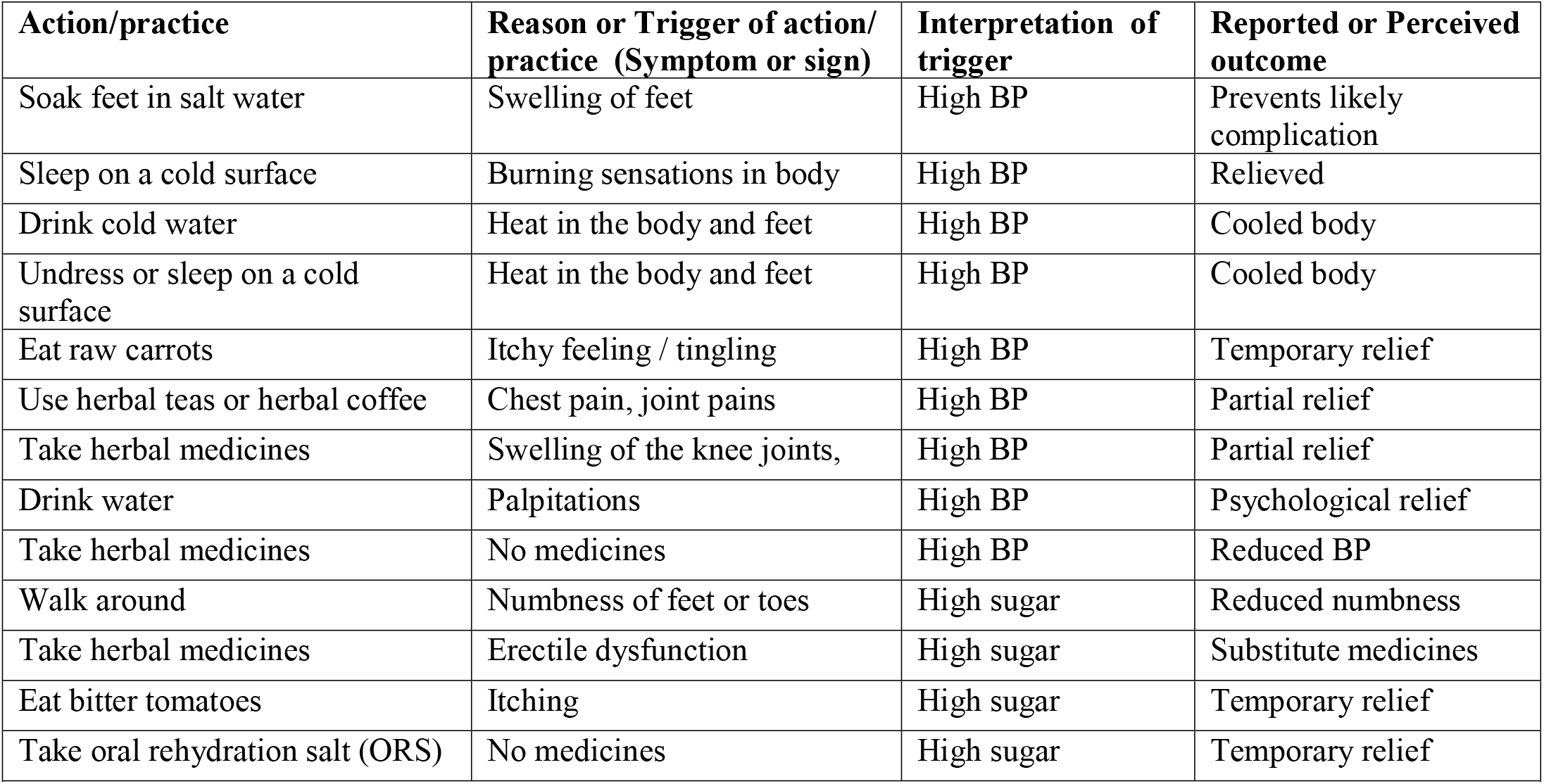
Practices in place of missed medicines (when conventional medicines are not available)

Patients also described using traditional medicines to ration prescribed medicines:

> *R: Sometimes when I fail to get all the medicines, I sparingly take them along with herbs*.
>
> ***I: Can you please clarify …***
>
> *R: If* [pharmacists] *have given me tablets, say, for 2 weeks and they told me to take two tablets thrice a day, what I normally do is to swallow two tablets twice a day, omiting these two tablets and push them forward. So instead of swallowing these two tablets during the day, I drink some herbal medicine during day. If I realize that my medicine is reducing and yet there are more days, I reduce and swallow one tablet in the morning and one tablet in the evening. So at around lunchtime, I drink my cup of herbal medicine which I have prepared*. (IDI-15, DM, Female, 36)

When patients were able to attain all their prescribed doses, they also saw traditional medicines as a useful *supplement* to prescribed medicines. Some patients used traditional medicines alongside conventional medicines to attain a desired health status, such as weight loss, or to cure their condition (Table 2B). For example, one patient described finding relief through an herbal concoction, despite its cost:

**Table 2B:**
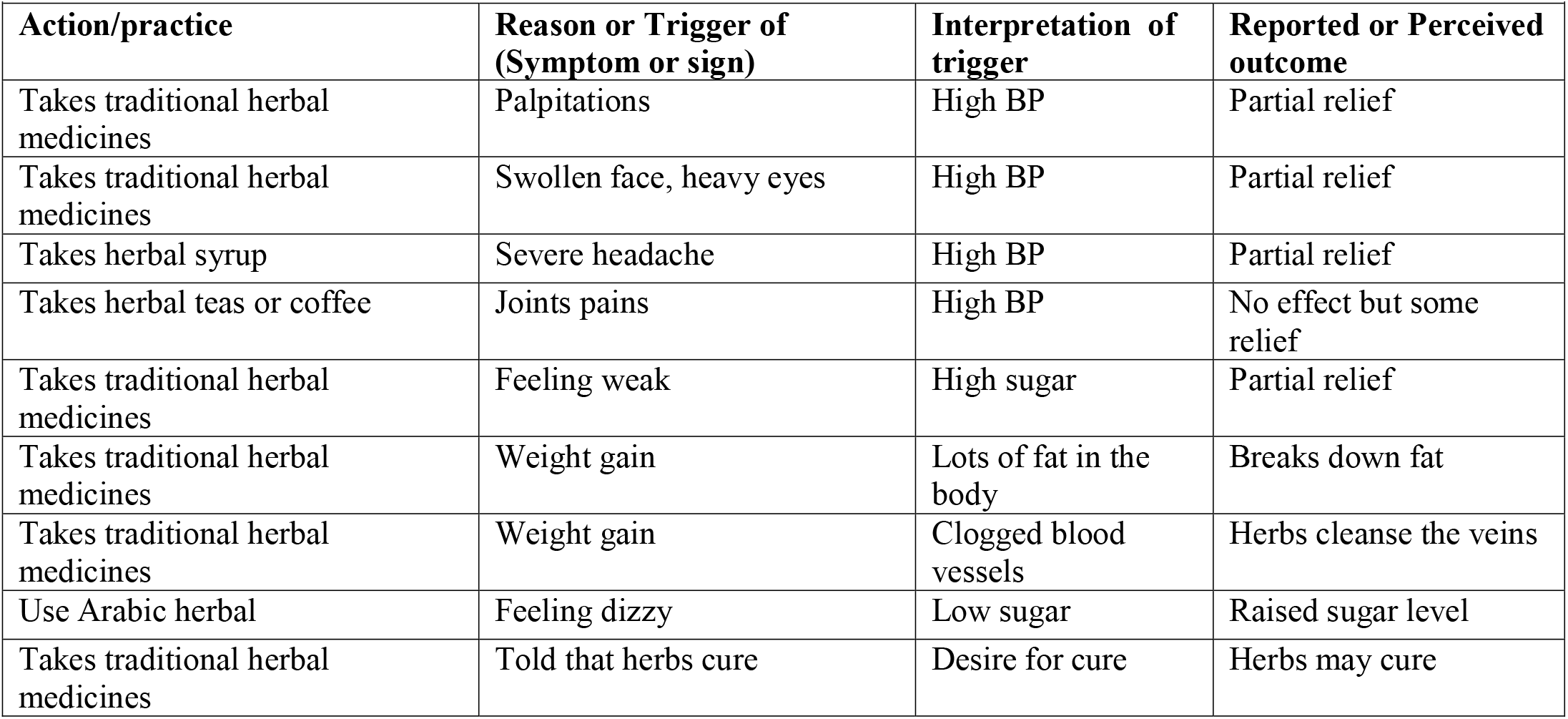
Use of traditional medicines alongside conventional medicines (when conventional medicines are available)

> *R: I have used some herbal medicine from a certain Hajjat, she insists that she heals diabetes and she really has medicine that heals diabetes. I even bought a jerry can worth UGX 80,000* [approximately $22 USD], *but doctors say that diabetes is incurable. … It really worked for me and gave me some relief. For instance, my sugar level was at 17 [mmol/l, equivalent to 306 mg/dl] and after drinking it reduced to 15 [270mg/dl], 12 [216mg/dl], 11 [198mg/dl] like that. I have not yet got enough money to buy more, she just gave me a mixed jerry can*.
>
> ***I: Did you use it because she said it heals diabetes?***
>
> *R: I used it because I heard her on radio saying she really heals diabetes. …, It is so helpful but she is expensive. I plan on buying it again when I get money, because it was of great help to me: it brought relief to me*. (IDI-11, DM_Male, 52)

### Patients adopted a wide range of vernacular practices to supplement treatments

Besides using medicines, patients restricted diet, sought ways to control stress, and improvised self-monitoring strategies in order to maintain their health. They also adopted a variety of self-care practices in response to specific symptoms, such as walking barefooted for relief when their feet were numb, soaking swollen feet in salt water, and massaging their bodies with warm water to relieve itching (Table 3). Concerning diet, one stated:

> *As a result of this disease, I decided to desist from soft drinks and all sweet things. I no longer take sugar, but life is not enjoyable. I eat mostly matooke [steam-cooked starchy bananas] and Irish potatoes. For cassava, I eat just a piece when I feel like. I mainly avoid fatty foods such as margarine and fatty meat. I however eat roasted meat*. (IDI-5, DM, Male, 65)

**Table 3:** Practices to supplement treatments.

| **Action/practice** | **Reason or Trigger of (Symptom or sign)** | **Interpretation of trigger** | **Reported or Perceived outcome** |
| --- | --- | --- | --- |
| Takes time off from people | Stressed or unhappy | Causes BP or sugar levels to increase | Feels better |
| Eats only traditional foods | Avoiding fatty foods | fatty foods unhealthy | to maintain health |
| Use anti-biotic | Feeling dizzy | High BP | Gets some relief |
| Avoids worrying | Lack of strength, feeling weak | High BP | Feels better |
| Eats immediately | Sudden hunger | High BP | Feels better |
| Stay away from foods (fasting) | Hearing the heart beats | High BP | To reduce weight |
| Takes a lot of water | Constant headache | High BP | Gets some relief |
| Takes a walk | Feeling weak | High BP | to improve blood circulation |
| Takes herbal medicines | Not feeling well | High BP | Reduced BP |
| Takes pain killer | Swelling of feet | High BP | Swelling reduced |
| Takes time off from others | Palpitations | High BP | Palpitations stopped |
| Use any pain killers | Severe headache | High BP | Gets relief |
| Walking | Breathing heavily | High BP | To sweat and reduce fat |
| Use pain killers | Numbness of feet or legs | High BP | Feels better |
| Drink lots of water | Frequent urination | High sugar | No effect |
| Eats food and takes a walk | Cloudy vision | High sugar | Felt better |
| Increase the medicine dosage | When the urine is yellow | High sugar | Gets some relief |
| Massage legs with warm water | Paralysis of feet and knees | High sugar | Gets some relief |
| Stretch legs morning and evening | Paralysis of feet and knees | High sugar | Perceived to help straighten blood vessels |
| Takes off time to sleep | General body weakness | High sugar | Gets some relief |
| Uses worm water | Itches | High sugar | Gets some relief |
| Walks barefooted on small stones | Numbness of feet | High sugar | Feels good |
| Drink lots of water | Feeling thirsty | High sugar | No-effect |
| Digging, walking and household chores | Main physical activities | Improve blood circulation | Body functions well |
| Eat sugar | Foaming in mouth | Low sugar | got better |
| Eat sweet bananas | Body weakness | Low sugar | Feels better |
| Take sugar | Temporarily unable to speak | Low sugar | Stabilized |
| Take something sweet (e.g., sweet bananas) | Cloudy vision | Low sugar | Gets better in one minute |
| Taste own urine | Anxious about sugar level | Sweet taste implies high sugar | Perceives the sugar level |

When patients with diabetes felt weak or experienced symptoms they linked to a low sugar level, many reported consuming sweet things to restore their blood sugar:

> *When I get a foam in the mouth to an extent that my tongue cannot move, I know that my sugar level is low. So I lick some sugar or dissolve it in water and I drink. It is my first medicine*.*… Even other diabetic patients have told me that I should take sugar the moment I notice that my sugar level has reduced*. (IDI-12, DM, Female, 78)

Because patients with diabetes were anxious to ascertain their sugar level but could not obtain conventional glucose tests, they improvised self-monitoring practices. For example, some patients reported tasting their urine to gauge sugar level:

> *R: Some of our colleagues [fellow support group patients] tell us that they are check their sugar levels using their urine and that’s what I currently do. If the urine is very sweet, then the sugar level is high but if it tastes like the salt of lake Katwe, then the level is low* (IDI-15, DM, Female, 36).

Patients saw these adopted practices as adjuncts to conventional treatments and diagnostic tests to which they lacked access, rather than as core practices for attaining or maintaining health.

### To mitigate uncertain access to medicines, patients sought social support for self-care

Patients attempted to mitigate uncertain access to conventional medicines by drawing on other resources in their social environment. In many instances, patients saw their children as their most important source of emotional and instrumental support to sustain self-care. In particular, children were a primary source of instrumental support, providing money for patients’ medicines, transport, and necessities at home:

> *The children buy the medicines and bring them. They can bring for me some medicines from Kampala where they stay. Sometimes children send money and I buy medicines*. (IDI-2, HTN, Female, 52).

Patients also looked to their spouses for social support. Some husbands looked to their wives for emotional support, food provision and preparation of meals. Wives tended to look to husbands solely for financial support. Patients counted on their children for financial support for treatment when financial support from the spouse dwindled:

> *They* [children] *help me. Because if I don’t have money for transport to come, they can mobilize for me*.
>
> ***I: Who are those?***
>
> *R: The children we produce, because I may fail and my husband also fails. So we resort to asking our children for money*. (IDI-10, HTN & DM, Female, 59).

Patients considered their children a final but frequently-called-upon resort for financial resources to sustain supplies of medicine.

Finally, patients reported receiving peer support through patient groups. Some patients said that they organized themselves into these groups for the purpose of mutual support; this was mostly reported among diabetes patients. In addition to emotional support, patients reported receiving instrumental support for transport and medicines from other patients as part of these mutual aid groups. For example, one patient described how patient groups organized and pooled resources for mutual support:

> *That group has helped more on the side of unity. Because we have the 1000UGX we collect every meeting but that money is for disaster preparedness such that in case one of us is not able to come due to transport challenges, he calls the leaders and they send him transport instead of staying home and dying… The group still helps me especially on the medicines and strips. We collect 3000 UGX per head so that in case government does not bring them, the doctors buy for us with our own money* (IDI-5, HTN & DM, Male, 65).

Second, patients reported receiving informational support about self-care practices from other patients in their group:

> *Our leaders in the group like that man* [a patient member of the support group], *sensitize us on how we can protect ourselves, feeding patterns, and the way we are supposed to behave. For instance, I do not have to be with a gloomy face, I have to be happy. So they come and give us health education* (IDI-10, HTN & DM, Female, 59).

Groups were a source of emotional support for patients who participated them. Patients reported watching out for one another and creating expectations for self-care, food relief and regular appointment attendance. A group leader explained:

> *We don’t allow anybody in our group to miss an appointment without calling us to let us know why. You must show up and in case you won’t come due to transport issues, you call us and we send you money so you report*.*…. Now, in case someone is ill and they don’t have money, should we let them die? So we decided to collect 1,000 UGX [<$1 USD] every time we come here. … We also lend to that person who doesn’t have what to eat*…, *And when you have lost a loved one, we give you condolence of 30,000 UGX [$8*.*1 USD]* (IDI-3, HTN & DM, Male, 76).

## Discussion

In this interview-based study of patients’ self-care for diabetes and hypertension in rural Uganda, we found that patients valued and used both conventional and traditional medicines, but preferred conventional medicines as their first line of resort. They also adopted vernacular practices to supplement conventionally recommended self-care practices. In response to inconsistent access to conventional medicines, patients sought support from family and peers.

Crucially, we found that patients relied on networks of social support to mitigate uncertain access to medicines and meet transport costs associated with care. Studies of chronic HIV care in Uganda demonstrate that families, friends and other significant persons can be important sources of similar support for adherence.^18, 19^ Similarly, a study in Nigeria identified families as the most accessible source of financial support for patients with hypertension and diabetes and found that this marginally impacted health outcomes.^20^ We add to this literature the notable finding that patients relied most on their children for support, rather than peers or spouses—even when they characterized their children as not financially stable. A lack of economic resources increases the importance of such social networks for accessing treatment.^6^ We identify a critical need for interventions to facilitate additional sources of instrumental and emotional support for patients.

We also found that many patients with diabetes participated in loosely organized patient groups. Patients pooled limited resources to provide test strips, transport support, emergency funds for medicines, and even food relief. While groups often spontaneously arose in clinic waiting rooms, they were not affiliated with the health facilities. This is consistent with an earlier study we conducted at Ugandan public hospitals, which found that informal diabetes support groups helped patients manage frequent stock-outs of medicines.^21^ Our study further affirms the centrality of group support as a social, emotional, and financial resource for managing health. This finding is particularly important because it suggests a viable local model for a cost-effective, patient-centered intervention to mitigate resource variability. Patient-driven groups can provide additional sources of non-family, peer support for all patients. Strengthening, and scaling up these groups could address gaps in social support, improve consistent access to essential medicines and supplies, facilitate effective self-care, and improve adherence to medicines. Further research should develop and evaluate interventions to expand and strengthen the patient peer support model for rural settings.

We also found that patients valued prescribed medicines and preferred them for disease management. Indeed, patients may even overestimate the capacity of these medicines; another study in rural Uganda showed that patients believed that consistent adherence to conventional medicines would cure hypertension and diabetes.^22^ However, patients also reported that their access to prescribed medicines was poor or inconsistent.

Insufficient medicine is the commonest reason for poor adherence to medication in Uganda.^5, 22^ In Uganda, public facilities are the primary source of free medicines for patients with NCDs. However, recent work has demonstrated that the majority of medicine doses prescribed for diabetes and cardiovascular disease at these facilities are not dispensed due to real or anticipated shortage.^23^ Therefore, patients must obtain their prescriptions elsewhere, if at all.^22, 24^ Future implementation research and policy should explore strategies to improve access to medicines, which could improve adherence to prescribed medicines and reduce reliance on non-recommended care practices.

Moreover, because they had no test supplies, patients in our study described improvising self-monitoring strategies. Chronic care in Uganda is persistently affected by poor availability of essential medicines and equipment.^25^ Similar gaps in NCD service provision have been reported in other developing countries like Malawi^26^, Ethiopia^27^, and Bangladesh.^28^ Future research should explore strategies to improve access not only to medicines but also to essential materials and equipment. Strategies to improve self-care in these settings must attend to the limited resources available for self-monitoring at home.

We also found that patients used traditional medicines to both mitigate the negative impact of inconsistent access and supplement conventional medicines. The use of traditional medicines to compensate for rationed prescription medication may imply that patients perceive traditional treatments to play a similar role as prescribed medicines, as has been reported in other studies Uganda and India.^21, 24, 29^ The use of traditional medicines is common in Uganda and influenced by several actors including traditional healers, peers, family members and media.^5, 21, 24^ However, patients in our study did not report ceasing conventional medicine in favor of traditional medicine, as others have found in south-western Uganda.^24^ Rather, patients in our study believed their prescribed medicines to be central to managing their condition. Additionally, patients in this study relied on biomedical monitoring to judge the effectiveness of the traditional medicines they consumed. This commitment to conventional medicine may reflect our recruitment strategy from conventional health facilities.

This work should be regarded within the context of some limitations. First, we recruited participants from health facilities. Therefore, our participants were more likely to have overcome certain barriers to accessing care and/or have stronger health-seeking behaviors than the general population. These findings may not reflect the experiences or practices of people who are not engaged in clinical care. Second, this qualitative study was conducted in just three rural settings. Future research should examine the patterns identified here in larger representative samples.

This work also has strengths. Our sample includes a balance of male and female patients with hypertension, diabetes, or both conditions. We also included participants from different types of health facilities, enhancing the external validity of our findings. Second, our analytic strategy involved a multi-disciplinary coding team, consensus meetings about the code structure, peer debriefing and reviews by all investigators, which facilitated both emergence of divergent interpretations and cross-coder reliability.

## Conclusion

Conventional medicines were a key resource for self-care among patients living with diabetes and hypertension in rural Uganda. However, patients faced uncertain and inconsistent access to conventional medicines, which affected their adherence to medicines and adoption of self-care practices. Patients therefore drew on networks of social support among their families and peers to reduce uncertainty and engage in mutual aid. Interventions to improve self-care and patient outcomes in such rural settings may be more effective if these interventions strengthen access to medicines and leverage existing support networks.

## Data Availability

All the required data is available in the manuscript

## Contributors

EV, CN, TLR, NH, JIS and MAH conceptualized the study. All authors made substantial contributions to the design and planning of the study. AKT the led data collection. AKT and MAH conducted the data analysis. All authors contributed to the interpretation of the data. AKT and MAH prepared the first drafts of the manuscript and all authors reviewed the initial manuscript draft. All authors reviewed and approved the final version of the manuscript.

## Acknowledgments

We are grateful to the following persons for their invaluable support: The District Health Officer of Nakaseke, the health facility administrators and who allowed us to conduct this research within the public sector facilities; all the NCD clinics’ staff and the research assistants who participated in data collection for this study. We are also grateful for the support from the Mixed methods training program.

## Appendix 1: In-depth interview guide

### Purpose statement

To explore patients’ descriptions of self-care practices regarding hypertension and diabetes.

### Introductory statement

We are gathered to discuss about our health. I would like you to share with me how you take care of yourself and what you do or avoid to do to keep in good health.

1. Could you please share with me how you got to know that you had hypertension or diabetes? (Probe: How did you know? What was happening in your life then/ symptoms?)
2. I would like to know your thoughts about living with diabetes or hypertension. How do you view this condition? (Follow-up question: Do you think you have enough information about this disease? Probe: what is the source of information?)
3. Let’s talk about the symptoms / signs that you face as patients with diabetes or hypertension. What are some of the symptoms? How do you recognize them? How do you respond / react to them when they occur? [*what meaning is placed on these practices or responses*]. (What about complications? What do you know about the complications that can affect you?)
4. Now let’s talk about the medical care you receive at this facility: How often are you supposed to attend this facility? Are you able to come for all appointments? (Probe: Why?)
5. We will talk more about the medicines you receive at the facility: Do you often get explanations for the medicines you receive? (Probe: frequency of obtaining prescribed medicines, what is done when medicines are not obtained at facility).
6. What are some of the challenges you face when attempting to adhere to medicines prescribed or even those you have obtained? (Probe: How easy or difficult to follow the recommended uptake of medicines give/ dosage?)
7. Apart from the medicines, is there anything else you do to be well, live happily or prevent adverse conditions? (Probe: what is done and why).
8. We will now talk more about our diet. Share with me your feeding practices ever since you were diagnosed with diabetes (Probe: what foods and drinks do you often eat/take? (Probe: Why the food/ drink, alcohol consumption, smoking and salt intake awareness and practice).
9. Let’s talk about exercises. Do you think these are necessary? Please explain. (Probe: Practices with regard to regular physical exercise importance of exercises, what they do or avoid to do to achieve physical activity?)
10. What are some of the other things (apart from diet and exercise) that the nurses or doctors have told you to do or to avoid? (Probe: Have you been able to perform them? Why not able to perform some recommendation? Why not able to perform other recommendation? Probe about monitoring of the disease, foot care, etc.)
11. What other issues or challenges do you experience when keeping up to maintain good health? (Probe: social /family support, societal influences, receipt of care from community health workers)
12. What do you think can be done to best improve your uptake of healthy behaviors recommended by the health workers?

